# Long Sleep Duration, Cognitive Performance, and the Moderating Role of Depression: A Cross-Sectional Analysis in the Framingham Heart Study

**DOI:** 10.1101/2024.12.02.24318350

**Authors:** Vanessa M. Young, Rebecca Bernal, Andree-Ann Baril, Joy Zeynoun, Crystal Wiedner, Carlos Gaona, Alexa Beiser, Antonio L. Teixeira, Arash Salardini, Matthew P. Pase, Jayandra Jung Himali, Sudha Seshadri

## Abstract

**INTRODUCTION:** We investigated whether depression modified the associations between sleep duration and cognitive performance.

**METHODS:** Multivariable linear regression models examined the associations between sleep duration and cognition in 1,853 dementia- and stroke-free participants from the Framingham Heart Study. Participants were categorized in four groups: no depressive symptoms, no antidepressants; depressive symptoms without antidepressants use; antidepressant use without depressive symptoms; both depressive symptoms and antidepressant use.

**RESULTS:** Long sleep was associated with reduced overall cognitive function. Strong associations between sleep duration and cognitive performance were found in individuals with depressive symptoms, regardless of antidepressant use. Weaker but significant effects were observed in those without depressive symptoms. No significant associations were observed in participants using antidepressants without depressive symptoms.

**DISCUSSION:** These findings provide new evidence that sleep duration may be a modifiable risk factor for cognitive decline, particularly in individuals with depressive symptoms. Future research should elucidate underlying mechanisms and temporal relationships.

**RESEARCH IN CONTEXT:** **Systematic review:** We conducted a systematic search on PubMed and Google Scholar for peer-reviewed articles using keyword combinations related to sleep, cognition, and depression. Existing evidence reveals mixed findings on the relationship between sleep duration and cognition, with limited studies examining the role of depression on this association.

**Interpretation:** We observed that only long sleep duration (≥9h) was associated with poorer global cognition, executive function, visuospatial memory, and verbal learning/memory. Depression moderated this association, showing stronger negative effects of long sleep on cognition in individuals with depressive symptoms, regardless of antidepressant use. This suggests that long sleep duration may serve as an early indicator or risk factor for cognitive decline in those with depressive symptoms.

**Future directions:** Longitudinal studies with objective and subjective sleep assessments across diverse populations are needed to clarify how depression and its treatment influence the relationship between sleep duration and cognitive decline and support clinical strategies.

**HIGHLIGHTS:**

1. Sleeping ≥ 9hours/night was associated with worse cognitive performance.
2. This association was stronger among those with depression.
3. Long sleepers were more likely to report symptoms of depression.
4. Sleep may be a modifiable risk for cognitive decline in people with depression.

## 1 BACKGROUND

Cognitive decline and dementia, particularly Alzheimer’s disease (AD), are pressing public health concerns, exacerbated by increasing life expectancy worldwide.^1^ Cognitive impairment interferes with activities of daily living, threatens independence and quality of life, and leads to considerable healthcare costs and caregiver burden.^2–4^ Despite recent advancements in treatment, the efficacy of the current anti-amyloid disease-modifying treatments is modest, and their use is associated with significant adverse effects.^5^ Therefore, it is important to continue to investigate possible prevention strategies based on modifiable risk factors and their potential interaction.^6^

There is an increasing recognition of the significance of sleep as a vital physiological process for brain health.^7,8^ Disturbances in sleep duration and patterns are present in both normal and pathological aging,^9–11^ and they contribute to an increased risk of cognitive deficits.^12^ The Global Council on Brain Health recommends 7 to 8 hours of nightly sleep for adults to preserve brain health.^13^ Several studies have suggested that both excessive and insufficient sleep relative to the prescribed duration are linked to impairments in cognitive domains, including memory, attention, and executive functioning.^18–21^ The exact nature of these relationships varies depending on the study and the characteristic of the sample. The largest such study, a pooled cohort study of more than 28,000 people in mid to late-life, found that sleep durations of less than four or more than ten hours was associated with faster cognitive decline; suggesting an inverted U curve relationship between sleep duration and cognition.^19^ In contrast, a cross-sectional study of healthy middle-aged subjects^20^ and a longitudinal^21^ study of older adults reported an association solely between short sleep duration (< 6 hours/night) and diminished cognition.^22^ Other cross-sectional,^23^ longitudinal^24^ and prospective^18^ studies have only found prolonged sleep (≥ 9 hours/night) correlated with cognitive deterioration. These inconsistencies in evidence hinder our understanding of the precise nature of the association between sleep duration and cognition.

Depression, a leading cause of disability globally and a modifiable risk factor for cognitive decline^6^, often co-occurs with sleep disorders.^25^ The association between sleep disorders and depression is well-established, with about 90% of people with depression reporting problems with sleep.^22^ Sleep disorders are believed to precede depression rather than the contrary,^26,27^ with insomnia being the most frequent sleep disorder associated with depressive symptoms.^28^ Depression has also been associated with an increased risk of cognitive deficits.^29^ The cognitive effects of depression appear to persist even after remission of depressive symptoms,^30^ contributing to a heightened vulnerability to dementia later in life.^31^ Depressive feelings frequently occur in dementia patients and may act as early markers of the condition.^32^

The complex relationship among sleep duration, cognitive function, and depression remains inadequately investigated. Further investigation into these often comorbid modifiable risk factors may lead to the development of more personalized prevention strategies. To address this knowledge gap, we used data from the Framingham Heart Study to cross-sectionally examine the associations of short, average, and long self-reported nighttime sleep duration with cognitive performance among dementia- and stroke-free-adults. We further explored whether depressive symptoms modified these associations. We hypothesized that (1) both long and short self-reported nighttime sleep durations would be associated with poorer cognitive performance, and (2) depressive symptoms would moderate these associations such that the relationship between sleep duration and cognitive performance would be stronger in individuals with depressive symptoms regardless of antidepressant usage, relative to those without depressive symptoms.

## 2 METHODS

### 2.1 Sample

The Framingham Heart Study (FHS) is a multi-generational, longitudinal study started in 1948. We included 1,853 participants from the FHS Third Generation, Omni 2, and New Offspring Spouse Cohorts. The study included dementia- and stroke-free participants who completed neuropsychological testing during their second examination cycle between 2008 and 2011. Figure 1 illustrates the sample selection. This study was conducted in accordance with the Declaration of Helsinki. The Institutional Review Board at Boston Medical Center approved the study protocol. All participants signed the informed consent.

### 2.2 Sleep Assessment

Habitual self-reported sleep duration was assessed during in-clinic visits, which occurred on average approximately 1.7 ± 1.0 years before the neuropsychological assessment in these cohorts. The participants responded to a prompt that asked them to list the number of hours they typically sleep. We classified sleep duration into three categories: short sleep duration (less than 6 hours), average sleep duration (between 6 and 9 hours) for the reference group, and long sleep duration (more than 9 hours).^33^

### 2.3 Cognitive Assessments

Trained neuropsychological raters assessed cognitive function using a comprehensive and standardized test battery.^34^ In this study, we included the following tests: Trail Making Test Parts A and B^35^, Visual Reproduction and Logical Memory Tests (sum of immediate and delayed recall scores) from the Weschler Memory Scale—Revised^36^, and the Similarities Test from the Wechsler Adult Intelligence Scale.^37^ The tests assessed attention, processing speed, and executive function; visuospatial memory; verbal learning and memory; and verbal reasoning, respectively. Trail Making Test Parts A and B were log-transformed and reversed, so that a higher score indicated better performance for all cognitive tests. A global cognitive score was derived from a principal component analysis,^38^ combining weighted loadings of the Similarities Test, Visual Reproduction, Logical Memory Test, and Trail Making Test Part B.

### 2.4 Measures of Depression

We used the 20-item Center for Epidemiologic Studies Depression Scale (CES-D)^39^ to assess depressive symptoms. We defined depression as either a score of ³ 16 on the CES-D or the use of antidepressant medications (anatomical therapeutic chemical [ATC] code N06A).^40^ We then categorized the participants into four groups: (1) controls – those not using antidepressants and without depressive symptoms (CES-D < 16); (2) those using antidepressants and without depressive symptoms (CES-D < 16); (3) those not using antidepressants with depressive symptoms (CES-D ≥ 16); and (4) those using antidepressants and with depressive symptoms (CES-D ≥ 16). The four-category classification affords clinical nuances in the relationship between depressive symptoms, treatment status, and cognitive outcomes that may be masked when using a binary classification (yes versus no) for overall prevalence estimates.

### 2.5 Covariates

The following covariates, potentially associated with sleep, depression, or cognitive function, were incorporated into two separate statistical models:

- Age (years) and age squared (years) at the time of neuropsychological testing.
- Education: high school or less, some college, or college degree.
- Sex: female or male.
- Cohort: Third Generation, Omni 2, and New Offspring Spouse Cohorts from FHS.
- Time interval between sleep duration questionnaires and neuropsychological testing.
- Hypertension was characterized according to the Seventh Report of the Joint National Committee on Prevention, Detection, Evaluation, and Treatment of High Blood Pressure, as stage I hypertension or worse, defined by a systolic blood pressure (SBP) of ≥140 mmHg, a diastolic blood pressure (DBP) of ≥90 mmHg, or the utilization of antihypertensive medication.^41^
- EDTA plasma aliquots were analyzed daily at the FHS laboratory to measure: (1) total cholesterol level (mg/dL), (2) high-density lipoprotein level (mg/dL)^42^, and (3) triglycerides (mg/dL).
- C-reactive protein (CRP) levels were categorized as low <1mg/L, average 1-3mg/L, and high >3mg/L, according to the American Heart Association guidelines.^43^
- Body mass index (BMI) calculated using weight in kilograms and divided by the square of height in meters (m^2^).
- *Apolipoprotein 4* allele carrier status (*APOE 4* status): those with at least one *4* allele (as determined by PCR) vs those with none.

### 2.6 Statistical Analyses

We used SAS V.9.4 (SAS Institute) to perform statistical analyses and considered statistical significance at *p* < 0.10 for test of interactions and *p* < 0.05 for all other tests. We treated self-reported sleep duration (short, average, and long sleep duration) and depression group (control, antidepressants usage/CES-D < 16, no antidepressants usage/CES-D ≥ 16, and antidepressants usage CES-D ≥ 16), as categorical variables.

First, we used multivariable linear regression models to examine the associations between the three sleep categories and global cognition. Secondary analysis assessed the associations between sleep duration categories and individual cognitive tasks. Model 1 was adjusted for age and age squared at neuropsychological testing, education, sex, time between sleep duration questionnaires and neuropsychological testing, and cohort. In Model 2, we additionally adjusted for hypertension, total cholesterol to HDL ratio, triglycerides, CRP level, BMI, and *APOE 4* status.

We explored effect modification by depression group for the associations between sleep duration and cognitive performance by adding sleep group by depression group interaction terms in Model 1. When a statistically significant interaction was observed, we stratified the association between sleep duration and cognitive performance by depression group.

### 2.7 Sensitivity Analysis

In the primary models between sleep duration and cognitive performance, we conducted a sensitivity analysis limiting the sample to middle aged and older adults (cut-off ≥ 45 years, following the age groups defined by the National Library of Medicine^44^) to examine whether the sleep duration-cognition association and potential effect modification by depression group remained consistent when excluding younger adults from the analysis.

## 3 RESULTS

### 3.1 Sample Characteristics

The sample consisted of 1,853 adults with an average age of 49.8 years (age range 27– 85). The sample was 42.7% male, 95.3% white, and 57.2% college educated. Most of the participants reported average sleep durations of 6-9 hours (68.9%), followed by short sleep (≤6 hours; 24.3%) and long sleep (≥9 hours; 6.8%). Depression, defined as CES-D ≥ 16 or the use of antidepressant medications, was present in 448 (24.2%) participants: 315 (17.0%) reported using antidepressants, while 198 (11.0) had a CES-D score of 16 or higher. The group with long sleep duration tended to have a higher proportion of depression (n = 55, 44.0%) compared to the short (n = 106, 23.5%) and average (n = 287, 22.5%) sleep groups. **Table 1** shows the overall sample’s characteristics, and **Table 2** shows the performance on domain-specific cognitive tests categorized by sleep duration.

**Table 1.**
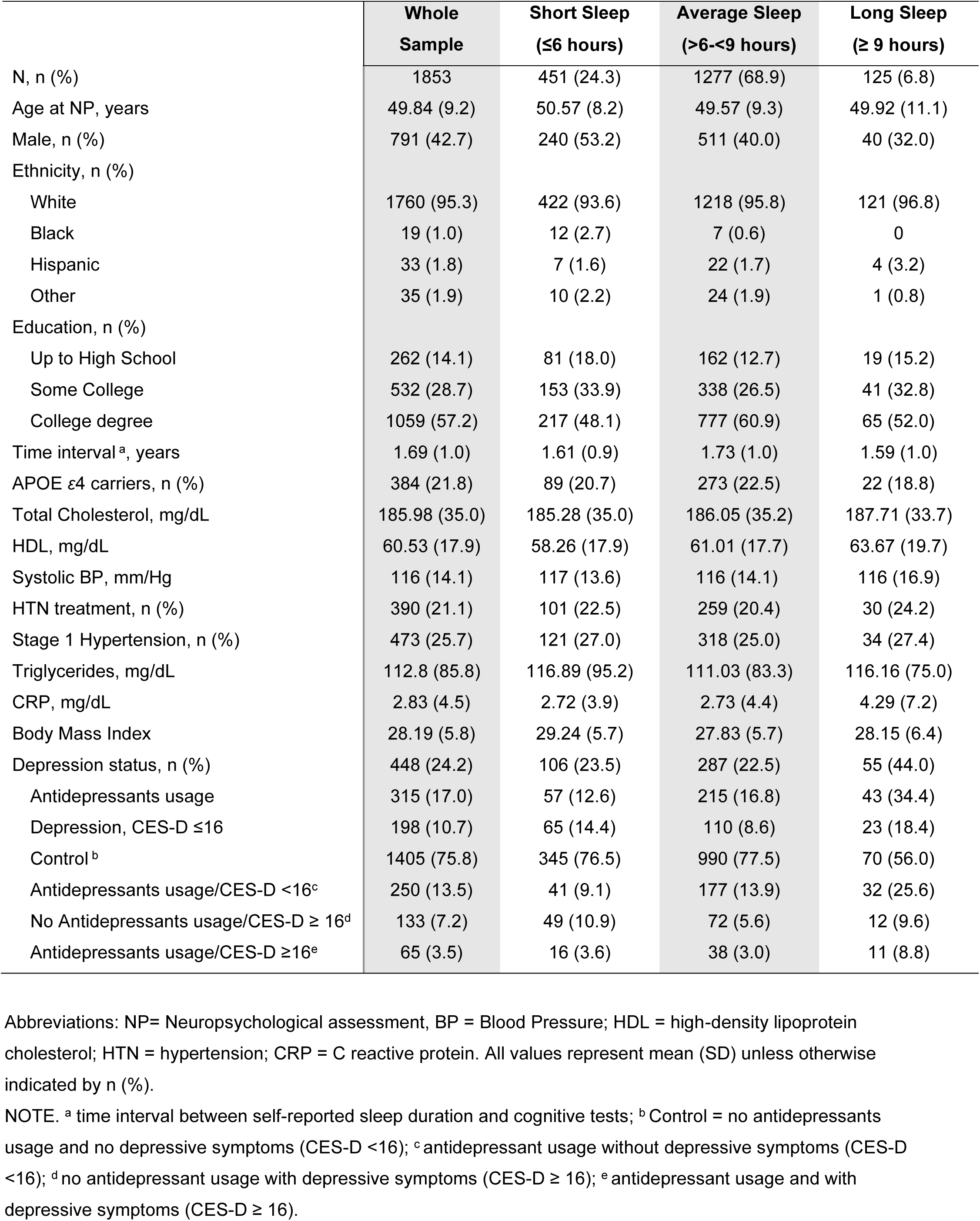
Sample characteristics.

|  | **Whole**<br>**Sample** | **Short Sleep**<br>**(≤6 hours)** | **Average Sleep**<br>**(>6-<9 hours)** | **Long Sleep**<br>**(≥ 9 hours)** |
| --- | --- | --- | --- | --- |
| N, n (%) | 1853 | 451 (24.3) | 1277 (68.9) | 125 (6.8) |
| Age at NP, years | 49.84 (9.2) | 50.57 (8.2) | 49.57 (9.3) | 49.92 (11.1) |
| Male, n (%) | 791 (42.7) | 240 (53.2) | 511 (40.0) | 40 (32.0) |
| Ethnicity, n (%) |  |  |  |  |
| White | 1760 (95.3) | 422 (93.6) | 1218 (95.8) | 121 (96.8) |
| Black | 19 (1.0) | 12 (2.7) | 7 (0.6) | 0 |
| Hispanic | 33 (1.8) | 7 (1.6) | 22 (1.7) | 4 (3.2) |
| Other | 35 (1.9) | 10 (2.2) | 24 (1.9) | 1 (0.8) |
| Education, n (%) |  |  |  |  |
| Up to High School | 262 (14.1) | 81 (18.0) | 162 (12.7) | 19 (15.2) |
| Some College | 532 (28.7) | 153 (33.9) | 338 (26.5) | 41 (32.8) |
| College degree | 1059 (57.2) | 217 (48.1) | 777 (60.9) | 65 (52.0) |
| Time interval<sup>a</sup>, years | 1.69 (1.0) | 1.61 (0.9) | 1.73 (1.0) | 1.59 (1.0) |
| APOE ε4 carriers, n (%) | 384 (21.8) | 89 (20.7) | 273 (22.5) | 22 (18.8) |
| Total Cholesterol, mg/dL | 185.98 (35.0) | 185.28 (35.0) | 186.05 (35.2) | 187.71 (33.7) |
| HDL, mg/dL | 60.53 (17.9) | 58.26 (17.9) | 61.01 (17.7) | 63.67 (19.7) |
| Systolic BP, mm/Hg | 116 (14.1) | 117 (13.6) | 116 (14.1) | 116 (16.9) |
| HTN treatment, n (%) | 390 (21.1) | 101 (22.5) | 259 (20.4) | 30 (24.2) |
| Stage 1 Hypertension, n (%) | 473 (25.7) | 121 (27.0) | 318 (25.0) | 34 (27.4) |
| Triglycerides, mg/dL | 112.8 (85.8) | 116.89 (95.2) | 111.03 (83.3) | 116.16 (75.0) |
| CRP, mg/dL | 2.83 (4.5) | 2.72 (3.9) | 2.73 (4.4) | 4.29 (7.2) |
| Body Mass Index | 28.19 (5.8) | 29.24 (5.7) | 27.83 (5.7) | 28.15 (6.4) |
| Depression status, n (%) | 448 (24.2) | 106 (23.5) | 287 (22.5) | 55 (44.0) |
| Antidepressants usage | 315 (17.0) | 57 (12.6) | 215 (16.8) | 43 (34.4) |
| Depression, CES-D ≤16 | 198 (10.7) | 65 (14.4) | 110 (8.6) | 23 (18.4) |
| Control<sup>b</sup> | 1405 (75.8) | 345 (76.5) | 990 (77.5) | 70 (56.0) |
| Antidepressants usage/CES-D <16<sup>c</sup> | 250 (13.5) | 41 (9.1) | 177 (13.9) | 32 (25.6) |
| No Antidepressants usage/CES-D ≥ 16<sup>d</sup> | 133 (7.2) | 49 (10.9) | 72 (5.6) | 12 (9.6) |
| Antidepressants usage/CES-D ≥16<sup>e</sup> | 65 (3.5) | 16 (3.6) | 38 (3.0) | 11 (8.8) |

Abbreviations: NP= Neuropsychological assessment, BP = Blood Pressure; HDL = high-density lipoprotein cholesterol; HTN = hypertension; CRP = C reactive protein. All values represent mean (SD) unless otherwise indicated by n (%).
NOTE. <sup>a</sup> time interval between self-reported sleep duration and cognitive tests; <sup>b</sup> Control = no antidepressants usage and no depressive symptoms (CES-D <16); <sup>c</sup> antidepressant usage without depressive symptoms (CES-D <16); <sup>d</sup> no antidepressant usage with depressive symptoms (CES-D ≥ 16); <sup>e</sup> antidepressant usage and with depressive symptoms (CES-D ≥ 16).

**Table 2.** Sample cognitive characteristics.

|  | **Whole Sample** | **Short Sleep**<br>**(≤6 hours)** | **Average Sleep**<br>**(>6-<9 hours)** | **Long Sleep**<br>**(≥ 9 hours)** |
| --- | --- | --- | --- | --- |
| N, n (%) | 1853 | 451 (24.3) | 1277 (68.9) | 125 (6.8) |
| Global Cognition, <sup>a</sup> median [Q1, Q3] | 0.45 [-0.12,0.98] | 0.32 [-0.19,0.91] | 0.49 [-0.06,1.01] | 0.15 [-0.49,0.88] |
| Trails Part A, min, median [Q1, Q3] | 0.40 [0.33,0.48] | 0.40 [0.33,0.48] | 0.40 [0.33,0.48] | 0.40 [0.32,0.55] |
| Trails Part B, min, median [Q1, Q3] | 0.97 [0.77,1.22] | 1.00 [0.77,1.30] | 0.95 [0.75,1.20] | 0.97 [0.80,1.33] |
| Visual Reproduction <sup>b</sup>, n correct | 17.99 (4.89) | 17.56 (5.09) | 18.31 (4.70) | 16.22 (5.56) |
| Logical Memories <sup>b</sup>, n correct | 24.46 (6.74) | 24.14 (6.59) | 24.71 (6.74) | 23.12 (7.12) |
| Similarities Test, n correct | 17.20 (3.15) | 17.08 (3.14) | 17.27 (3.17) | 16.95 (3.01) |

Abbreviations: Min = minutes.
NOTE. <sup>a</sup> Weighted score units; <sup>b</sup> Sum of immediate and delayed recall scores. All values represent mean (SD) unless otherwise indicated by median [Q1,Q3].

When examining sleep duration across the four depression categories (see **Table 3**), those using antidepressants and with depressive symptoms (CES-D ≥ 16) reported the highest proportion of long sleep (n = 11; 16.9%), followed by those using antidepressants but without depressive symptoms (n = 32; 12.8%). In contrast, the control group (no antidepressants usage and without depressive symptoms) had the lowest proportion of long sleepers (n = 70; 5.0%).

**Table 3.**
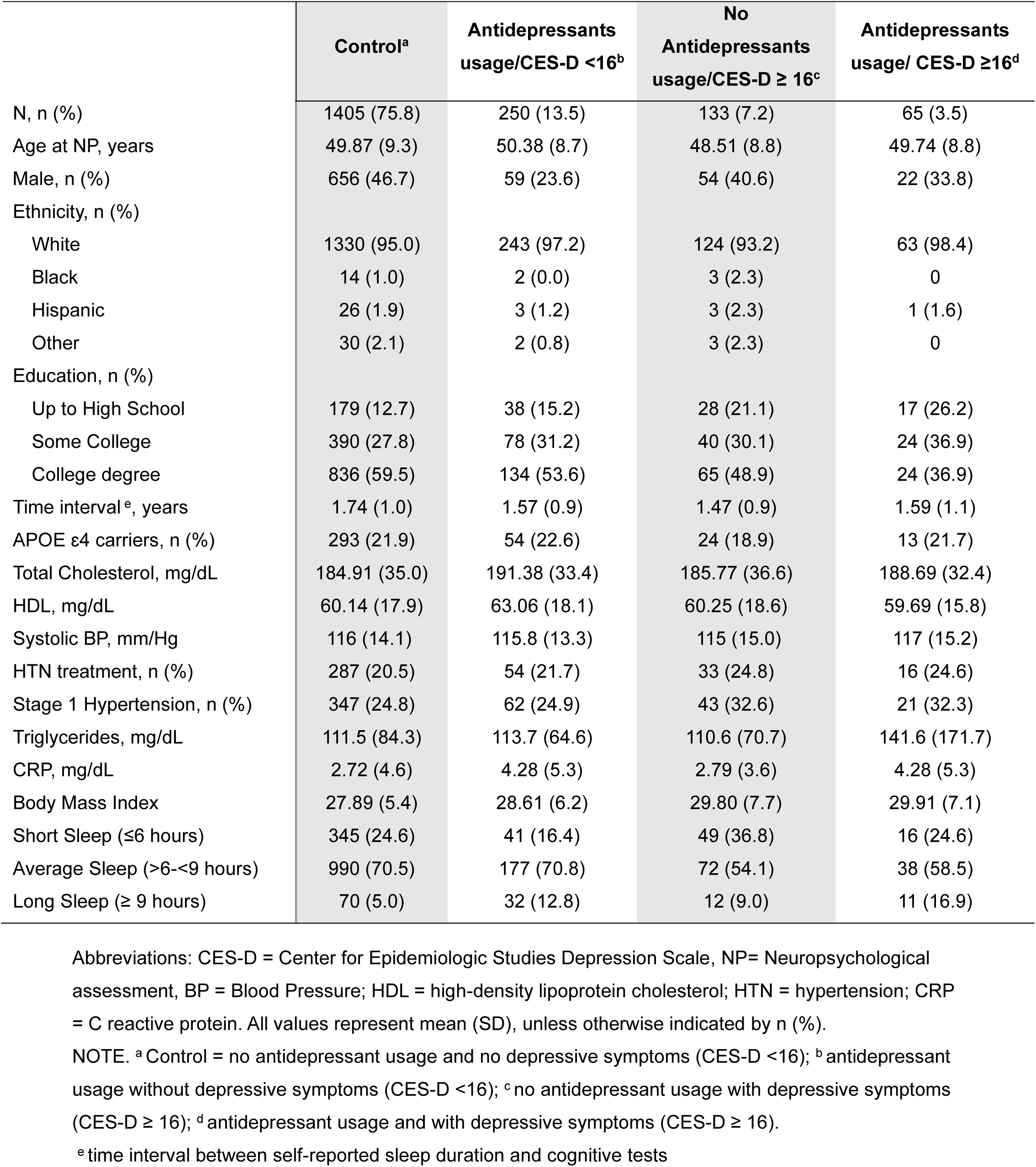
Sample characteristics by depression group.

|  | **Control<sup>a</sup>** | **Antidepressants**<br>**usage/CES-D <16<sup>b</sup>** | **No**<br>**Antidepressants**<br>**usage/CES-D ≥ 16<sup>c</sup>** | **Antidepressants**<br>**usage/ CES-D ≥16<sup>d</sup>** |
| --- | --- | --- | --- | --- |
| N, n (%) | 1405 (75.8) | 250 (13.5) | 133 (7.2) | 65 (3.5) |
| Age at NP, years | 49.87 (9.3) | 50.38 (8.7) | 48.51 (8.8) | 49.74 (8.8) |
| Male, n (%) | 656 (46.7) | 59 (23.6) | 54 (40.6) | 22 (33.8) |
| Ethnicity, n (%) |  |  |  |  |
| White | 1330 (95.0) | 243 (97.2) | 124 (93.2) | 63 (98.4) |
| Black | 14 (1.0) | 2 (0.0) | 3 (2.3) | 0 |
| Hispanic | 26 (1.9) | 3 (1.2) | 3 (2.3) | 1 (1.6) |
| Other | 30 (2.1) | 2 (0.8) | 3 (2.3) | 0 |
| Education, n (%) |  |  |  |  |
| Up to High School | 179 (12.7) | 38 (15.2) | 28 (21.1) | 17 (26.2) |
| Some College | 390 (27.8) | 78 (31.2) | 40 (30.1) | 24 (36.9) |
| College degree | 836 (59.5) | 134 (53.6) | 65 (48.9) | 24 (36.9) |
| Time interval <sup>e</sup>, years | 1.74 (1.0) | 1.57 (0.9) | 1.47 (0.9) | 1.59 (1.1) |
| APOE ε4 carriers, n (%) | 293 (21.9) | 54 (22.6) | 24 (18.9) | 13 (21.7) |
| Total Cholesterol, mg/dL | 184.91 (35.0) | 191.38 (33.4) | 185.77 (36.6) | 188.69 (32.4) |
| HDL, mg/dL | 60.14 (17.9) | 63.06 (18.1) | 60.25 (18.6) | 59.69 (15.8) |
| Systolic BP, mm/Hg | 116 (14.1) | 115.8 (13.3) | 115 (15.0) | 117 (15.2) |
| HTN treatment, n (%) | 287 (20.5) | 54 (21.7) | 33 (24.8) | 16 (24.6) |
| Stage 1 Hypertension, n (%) | 347 (24.8) | 62 (24.9) | 43 (32.6) | 21 (32.3) |
| Triglycerides, mg/dL | 111.5 (84.3) | 113.7 (64.6) | 110.6 (70.7) | 141.6 (171.7) |
| CRP, mg/dL | 2.72 (4.6) | 4.28 (5.3) | 2.79 (3.6) | 4.28 (5.3) |
| Body Mass Index | 27.89 (5.4) | 28.61 (6.2) | 29.80 (7.7) | 29.91 (7.1) |
| Short Sleep (≤6 hours) | 345 (24.6) | 41 (16.4) | 49 (36.8) | 16 (24.6) |
| Average Sleep (>6-<9 hours) | 990 (70.5) | 177 (70.8) | 72 (54.1) | 38 (58.5) |
| Long Sleep (≥ 9 hours) | 70 (5.0) | 32 (12.8) | 12 (9.0) | 11 (16.9) |

Abbreviations: CES-D = Center for Epidemiologic Studies Depression Scale, NP= Neuropsychological assessment, BP = Blood Pressure; HDL = high-density lipoprotein cholesterol; HTN = hypertension; CRP = C reactive protein. All values represent mean (SD), unless otherwise indicated by n (%).
NOTE. <sup>a</sup> Control = no antidepressant usage and no depressive symptoms (CES-D <16); <sup>b</sup> antidepressant usage without depressive symptoms (CES-D <16); <sup>c</sup> no antidepressant usage with depressive symptoms (CES-D ≥ 16); <sup>d</sup> antidepressant usage and with depressive symptoms (CES-D ≥ 16).
<sup>e</sup> time interval between self-reported sleep duration and cognitive tests

**Table 4.** Cognitive characteristics by depression group.

|  | **Control <sup>a</sup>** | **Antidepressants usage**<br>**/CES-D <16 <sup>b</sup>** | **No Antidepressants**<br>**usage /CES-D ≥ 16 <sup>c</sup>** | **Antidepressants usage**<br>**/CES-D ≥16 <sup>d</sup>** |
| --- | --- | --- | --- | --- |
| N, n (%) | 1405 (75.82) | 250 (13.49) | 133 (7.18) | 65 (3.51) |
| Global Cognition <sup>e</sup>, median [Q1, Q3] | 0.47 [-0.09, 0.99] | 0.41 [-0.19, 0.91] | 0.42 [-0.33, 0.96] | 0.12 [-0.38, 0.75] |
| Trails Part A, min, median [Q1, Q3] | 0.40 [0.33, 0.48] | 0.40 [0.33, 0.48] | 0.42 [0.33, 0.50] | 0.42 [0.32, 0.52] |
| Trails Part B, min, median [Q1, Q3] | 0.95 [0.75, 1.22] | 0.98 [0.78, 1.20] | 0.95 [0.75, 1.27] | 1.12 [0.88, 1.45] |
| Visual Reproduction <sup>f</sup>, n correct | 18.21 (4.81) | 17.44 (4.64) | 17.70 (5.53) | 15.88 (5.56) |
| Logical Memories <sup>f</sup>, n correct | 24.44 (6.70) | 24.99 (7.03) | 24.17 (6.53) | 23.46 (7.04) |
| Similarities Test, n correct | 17.26 (3.10) | 17.12 (2.97) | 16.87 (3.42) | 16.94 (4.10) |

Abbreviations: CES-D = Center for Epidemiologic Studies Depression Scale, Min = minutes.
NOTE. <sup>a</sup> Control = no antidepressants usage and no depressive symptoms (CES-D <16); <sup>b</sup> antidepressant usage without depressive symptoms (CES-D <16); <sup>c</sup> no antidepressant usage with depressive symptoms (CES-D ≥ 16); <sup>d</sup> antidepressant usage and with depressive symptoms (CES-D ≥ 16).
<sup>e</sup> weighted score units; <sup>f</sup> Sum of immediate and delayed recall scores combined). All values represent mean (SD) unless otherwise indicated by median [Q1, Q3]

### 3.2 Association between Sleep and Cognition

#### 3.2.1 Long Sleep Duration was Associated with Global Cognition

**Table 5** presents the association between short and long sleep durations and global cognitive performance, compared to average sleep duration (> 6 or < 9 hours vs. 6-9 hours). Long sleep duration (≥ 9 hours) had a statistically significant association with poorer global cognition (β *± SE=* −0.25 ± 0.07, *p* < 0.001), as measured by the global cognitive score, relative to those with average sleep duration. The association between short sleep duration (≤ 6 hours) and global cognition did not reach statistical significance (−0.03 ± 0.04, *p* = 0.465). In Model 2, the pattern of associations between long sleep duration and global cognition remained consistent with Model 1.

**Table 5.** Association between short and long sleep duration and cognitive scores.

| Cognition | Model | Average<br>(>6-<9 hours) | Sleep Duration Categories |  |  |  |
| --- | --- | --- | --- | --- | --- | --- |
|  |  |  | Short Sleep<br>(≤6 hours) |  | Long Sleep<br>(≥ 9 hours) |  |
|  |  |  | β±SE | p | β±SE | p |
| Global Cognition | 1 | REF | -0.03±0.04 | 0.465 | **-0.25±0.07** | **<0.001** |
|  | 2 | REF | -0.04±0.04 | 0.280 | **-0.27±0.07** | **<0.001** |
| Trails Part A | 1 | REF | -0.001±0.02 | 0.959 | -0.03±0.03 | 0.205 |
|  | 2 | REF | -0.01±0.02 | 0.550 | -0.03±0.03 | 0.279 |
| Trails Part B | 1 | REF | -0.02±0.02 | 0.235 | **-0.09±0.03** | **0.009** |
|  | 2 | REF | -0.04±0.02 | 0.067 | **-0.07±0.03** | **0.041** |
| Visual Reproduction | 1 | REF | -0.41±0.25 | 0.104 | **-1.80±0.42** | **<0.001** |
|  | 2 | REF | -0.45±0.26 | 0.081 | **-1.76±0.44** | **<0.001** |
| Logical Memory | 1 | REF | 0.25±0.35 | 0.480 | **-1.50±0.60** | **0.013** |
|  | 2 | REF | 0.22±0.36 | 0.538 | **-2.16±0.62** | **<0.001** |
| Similarities Test | 1 | REF | 0.01±0.17 | 0.954 | -0.06±0.28 | 0.839 |
|  | 2 | REF | 0.002±0.17 | 0.990 | -0.11±0.29 | 0.701 |

NOTE. Model 1 was adjusted for age and age squared at neuropsychological testing, education, sex, time between sleep duration questionnaires and neuropsychological testing, and cohort. Model 2 was further adjusted for hypertension, total cholesterol level to HDL ratio, triglycerides, C reactive protein level, body mass index, and APOE ε4 status.

#### 3.2.2 Long Sleep Duration was Associated with Task-specific Cognitive Domains

Long sleep duration (≥ 9 hours) was associated with significantly worse performance on Trail Making Test Part B (−0.09 ± 0.03, *p* = 0.009), Visual Reproduction (−1.80 ± 0.42, *p* < 0.001), and Logical Memory (−1.50 ± 0.60, *p* = 0.013) compared to the average sleep duration group. Long sleep was not associated with Trail Making Test Part A or the Similarities Test. Short sleep was not associated with any cognitive tasks. The results remained consistent in the fully adjusted model (see **Table 5**).

#### 3.2.3 Sleep Duration Interacts with Depression in its Association with Cognition

We tested statistical interactions to determine whether depression groups modified the relationships between sleep duration categories and cognitive performance. We observed significant interactions indicating effects on global cognitive score (p = 0.028), Trail Making Test Part B (p = 0.012), Visual Reproduction (p = 0.064), and Similarities test (p = 0.027).

As shown in **Table 6**, in the control group (no antidepressants, CES-D < 16), long sleep was associated with poorer global cognition (−0.18 ± 0.09, *p* = 0.044) and visual reproduction (−1.30 ± 0.55, *p* = 0.019), while short sleep showed no significant associations with any cognitive measure. In those with depressive symptoms (CES-D ≥ 16) not using antidepressants, long sleep was associated with poorer performance across multiple domains: global cognition (−0.60 ± 0.26, *p* = 0.024), Trails Part B (−0.29 ± 0.13, *p* = 0.023), and visual reproduction (−3.70 ± 1.70, *p* = 0.031). Short sleep was not significantly associated with cognitive measures in this group. Among those using antidepressants with depressive symptoms (CES-D ≥ 16), long sleep was associated with poorer global cognition (−0.74 ± 0.30, *p* = 0.017) and visual reproduction (−4.30 ± 1.89, *p* = 0.027). We observed no significant associations between short sleep and cognitive performance in individual using antidepressants but with depressive symptoms. Finally, among participants using antidepressants but without depressive symptoms (CES-D < 16), neither short nor long sleep duration was significantly associated with cognitive performance.

**Table 6.**
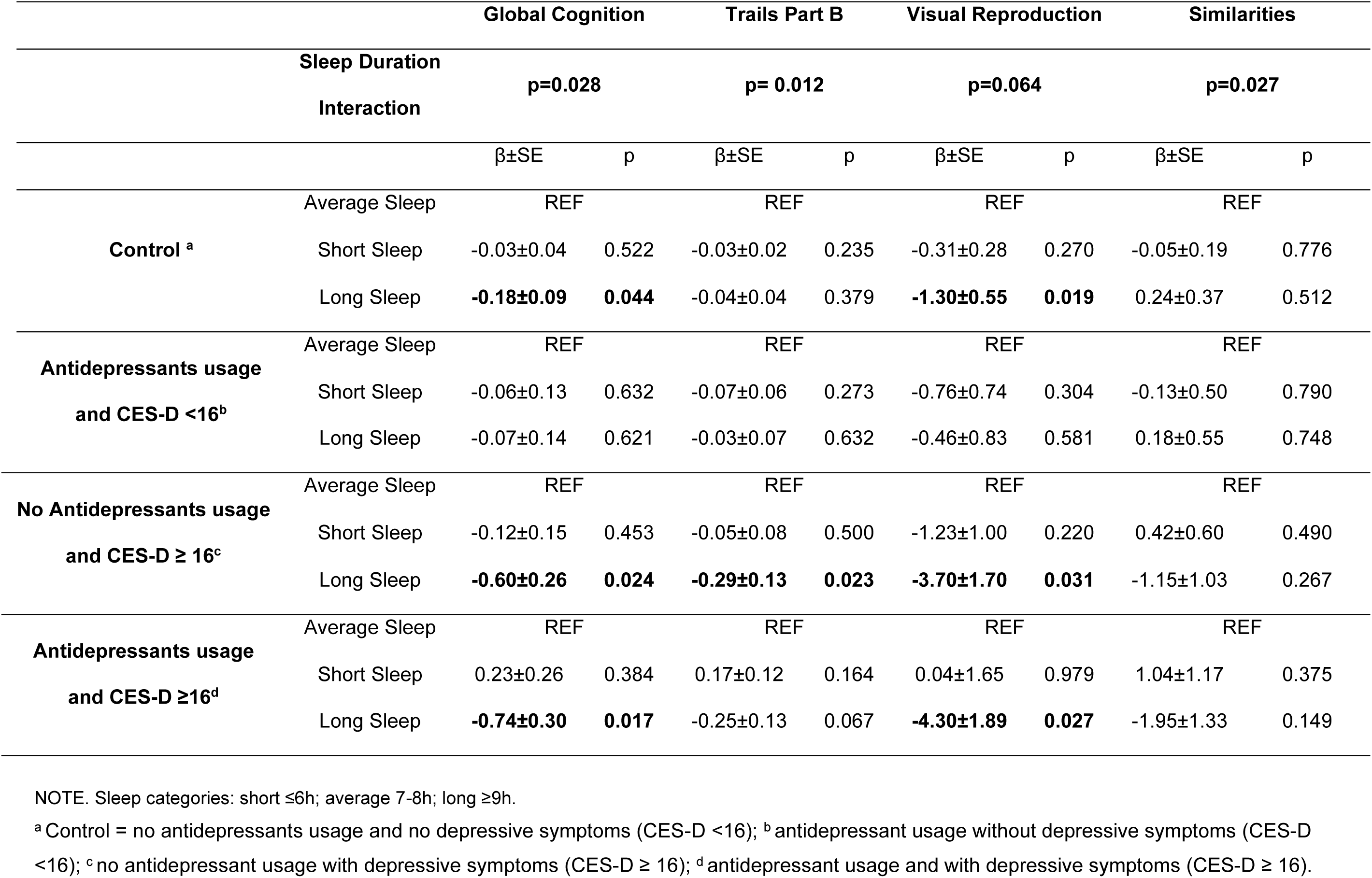
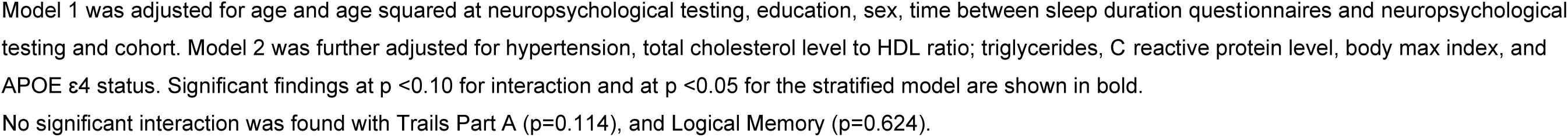
Effect modification by depression group on the association between self-reported sleep duration and cognitive scores.

|  |  | Global Cognition |  | Trails Part B |  | Visual Reproduction |  | Similarities |  |
| --- | --- | --- | --- | --- | --- | --- | --- | --- | --- |
| Sleep Duration Interaction |  | p=0.028 |  | p= 0.012 |  | p=0.064 |  | p=0.027 |  |
|  |  | β±SE | p | β±SE | p | β±SE | p | β±SE | p |
| **Control <sup>a</sup>** | Average Sleep | REF |  | REF |  | REF |  | REF |  |
|  | Short Sleep | -0.03±0.04 | 0.522 | -0.03±0.02 | 0.235 | -0.31±0.28 | 0.270 | -0.05±0.19 | 0.776 |
|  | Long Sleep | **-0.18±0.09** | **0.044** | -0.04±0.04 | 0.379 | **-1.30±0.55** | **0.019** | 0.24±0.37 | 0.512 |
| **Antidepressants usage and CES-D <16<sup>b</sup>** | Average Sleep | REF |  | REF |  | REF |  | REF |  |
|  | Short Sleep | -0.06±0.13 | 0.632 | -0.07±0.06 | 0.273 | -0.76±0.74 | 0.304 | -0.13±0.50 | 0.790 |
|  | Long Sleep | -0.07±0.14 | 0.621 | -0.03±0.07 | 0.632 | -0.46±0.83 | 0.581 | 0.18±0.55 | 0.748 |
| **No Antidepressants usage and CES-D ≥ 16<sup>c</sup>** | Average Sleep | REF |  | REF |  | REF |  | REF |  |
|  | Short Sleep | -0.12±0.15 | 0.453 | -0.05±0.08 | 0.500 | -1.23±1.00 | 0.220 | 0.42±0.60 | 0.490 |
|  | Long Sleep | **-0.60±0.26** | **0.024** | **-0.29±0.13** | **0.023** | **-3.70±1.70** | **0.031** | -1.15±1.03 | 0.267 |
| **Antidepressants usage and CES-D ≥16<sup>d</sup>** | Average Sleep | REF |  | REF |  | REF |  | REF |  |
|  | Short Sleep | 0.23±0.26 | 0.384 | 0.17±0.12 | 0.164 | 0.04±1.65 | 0.979 | 1.04±1.17 | 0.375 |
|  | Long Sleep | **-0.74±0.30** | **0.017** | -0.25±0.13 | 0.067 | **-4.30±1.89** | **0.027** | -1.95±1.33 | 0.149 |

NOTE. Sleep categories: short ≤6h; average 7-8h; long ≥9h.
<sup>a</sup> Control = no antidepressants usage and no depressive symptoms (CES-D <16); <sup>b</sup> antidepressant usage without depressive symptoms (CES-D <16); <sup>c</sup> no antidepressant usage with depressive symptoms (CES-D ≥ 16); <sup>d</sup> antidepressant usage and with depressive symptoms (CES-D ≥ 16).
Model 1 was adjusted for age and age squared at neuropsychological testing, education, sex, time between sleep duration questionnaires and neuropsychological testing and cohort. Model 2 was further adjusted for hypertension, total cholesterol level to HDL ratio; triglycerides, C reactive protein level, body mass index, and APOE $\epsilon$ 4 status. Significant findings at $p < 0.10$ for interaction and at $p < 0.05$ for the stratified model are shown in bold.
No significant interaction was found with Trails Part A ( $p=0.114$ ), and Logical Memory ( $p=0.624$ ).

#### 3.2.4 Sensitivity Analysis

In a sensitivity analysis including middle age and older adults (cut-off ≥ 45 years; see **Supplementary Tables 1** and **2** for sample characteristics and cognitive characteristics by age group), the pattern of associations between sleep duration and cognitive performance was largely consistent with the full sample results as shown in Supplementary **Table 3**. Significant interactions indicating effect modification by depression group were observed in the associations between sleep duration and global cognitive score (*p* = 0.009), Trails Part B (*p* = 0.038), Visual Reproduction (*p* = 0.050), and Similarities Test (*p* = 0.006). The most pronounced effects were seen in those using antidepressants and with depressive symptoms (CES-D ≥ 16). Relative to average sleep duration, long sleep duration was associated with significantly worse performance in all four cognitive measures: global cognition (−1.10 ± 0.35, *p* = 0.004), Trails Part B (−0.36 ± 0.15, *p* = 0.023), Visual Reproduction (−6.59 ± 2.24, *p* = 0.005), and Similarities Test (−3.80 ± 1.69, *p* = 0.030). In contrast, for the control group, only Visual Reproduction showed a significant negative association with long sleep duration (−1.43 ± 0.68, *p* = 0.037). No significant associations were observed among those using antidepressants and no depressive symptoms (CES-D < 16) and those not using antidepressants with depressive symptoms (CES-D ≥ 16). See Supplementary **Table 4**.

## 4 DISCUSSION

In this study, we investigated the association between self-reported sleep duration and cognition in the FHS cohorts and examined whether this association was modified by depression. Relative to average sleep duration, we found that long but not short sleep duration was associated with poorer performance in global cognition, executive function, visuospatial memory, and verbal learning/memory performance. Further, we found the strongest negative associations among individuals with depressive symptoms, regardless of antidepressant usage status (yes versus no).

Our initial hypothesis posited that both long and short sleep durations correlate with poorer cognitive function, as the balance of evidence points to a U-shaped association between sleep duration and cognition.^14–18^ Our findings partially support this hypothesis, corroborating the association between long sleep duration, worse global cognition, and poorer performance in multiple cognitive domains.^14–18^ Contrary to our expectations, we did not find a statistically significant relationship between short sleep duration and cognitive performance, with findings remaining consistent in the sensitivity analysis, including middle-aged and older adults (≥45 years) only. These results diverge from some studies that have identified short sleep as a risk factor for cognitive impairment,^14–18^ while aligning with others who reported a significant association between long sleep duration and poorer cognition.^23,24^

When exploring the role of depression in the relationship between sleep duration and cognition, we found that, in individuals with depressive symptoms (CES-D ≥ 16), associations between long sleep and cognition were more pronounced relative to controls. Specifically, in those with no prescribed antidepressants but with depressive symptoms (CES-D ≥ 16), long sleep was significantly associated with poorer performance in global cognition, executive function, and visuospatial memory. In those using antidepressants and with depressive symptoms (CES-D ≥ 16), long sleep was associated with worse global cognitive score and visuospatial memory. Conversely, no associations were found in those using antidepressants and potentially remitted depression (CES-D < 16), suggesting that effective treatment may help normalize sleep patterns and, as a consequence, improve cognitive function in some individuals with depression.^45^ These findings contrast with a recent study by Wang et al.,^46^ who found that long sleep duration was associated with better cognitive scores (TICS-10, word recall, figure drawing) among those with depressive symptoms (CES-D 10 ≥ 10) among 12,589 participants aged 45 and older from the China Health and Retirement Longitudinal Study (CHARLS). While both studies highlight the complex relationship between sleep, cognition, and depression, several factors could account for the divergent results. Differences in study populations, with Wang et al. concentrating on a Chinese cohort and our research predominantly involving White individuals residing in the U.S., suggest possible environmental, cultural, and/or ethnic differences in sleep patterns and the manifestation of depression symptoms.^47^ In addition, the cognitive assessments varied substantially, with our study using a comprehensive neuropsychological battery, potentially capturing subtle cognitive deficits associated with long sleep duration that could have been missed by the shorter screening measures used by Wang and colleagues.^46^ Further research is required to shed light on these relationships across diverse populations.

The reciprocal association between sleep disturbances and depression may create a cycle in which disrupted sleep exacerbates depressed symptoms, which in turn influences sleep quality.^48^ Sleep disturbance may increase vulnerability to depression by altering neural sensitivity to inflammation^49^ and disrupting neurotransmitter systems involved in mood regulation, such as serotonin and dopamine.^50^ Conversely, depressive symptoms can lead to sleep disruptions due to factors like worrying and difficulty coping with stress.^51^ Chronic stress and associated physiological changes, such as inflammation and dysregulation of the hypothalamic-pituitary-adrenal (HPA) axis, can further exacerbate depression and cognitive impairment.^52,53^ In those with depression, the HPA axis is usually hyperactive, potentially leading to fragmented sleep.^54,55^ The poorer cognitive performance observed in long sleepers may partly be attributed to lighter, fragmented sleep associated with depression characterized by frequent awakenings and less time spent in restorative, consolidated sleep stages. Slow-wave sleep (SWS) is crucial in cognitive function and brain maintenance.^56–58^ During SWS, the brain engages in a clearance process in which the glymphatic system eliminates metabolic waste products such as beta-amyloid and tau proteins.^56–58^ Fragmented sleep can suppress or reduce the duration of SWS,^59^ impeding this clearance mechanisms and potentially facilitating the buildup of neurotoxic proteins leading to cognitive deterioration. Future research with using objective measures such as polysomnography and actigraphy can provide insights into sleep architecture and fragmentation and help clarify the mechanisms which link long sleep duration to cognition.

Studies have suggested that long sleep duration may be an early and potentially reversible biological marker of neurodegeneration and a risk factor for dementia onset later in life.^60,61^ In our cross-sectional analysis, we observed that long sleep duration was associated with lower performance in specific cognitive domains, particularly executive function and visuospatial memory. These associations were present across our full age range, including adults younger than 45 years, and may reflect broader relationships between sleep duration and cognition rather than neurodegenerative processes specifically. Longitudinal studies integrating multimodal biomarker assessments of sleep, neuropsychiatric symptoms, and AD pathology are needed to better understand these relationships and to identify opportunities for therapeutic intervention.

Finally, significant associations between long sleep duration and poorer cognitive performance were also found in the control group (not using antidepressants and without depressive symptoms). The control group had a much larger sample size compared to the other three depression groups, which increases statistical power and the likelihood of detecting even small effects. The effect sizes in the control group, however, were smaller than in the groups with depressive symptoms. Hence, while the associations were statistically significant, they may not be clinically significant. Nonetheless, these associations highlight how the relationship between sleep duration and cognitive function is not exclusive to those with depressive symptoms. Underlying mechanisms operating independently of depression, such as sleep architecture, circadian rhythms^62^, or subclinical depressive symptoms^63^, may influence the relationship sleep duration and cognition. Earlier identification of changes in sleep duration and cognitive abilities could facilitate timely prevention and intervention, particularly against accelerated cognitive decline. A multidisciplinary care model involving sleep specialists, neuropsychologists, and mental health professionals may be valuable in enhancing early detection and intervention strategies. Further research is needed to determine which specific sleep interventions, such as cognitive behavioral therapy for insomnia (CBT-I), sleep hygiene education, or light therapy, might be most effective and feasible to implement in clinical practice for people with depression and to determine whether existing interventions are effective or require adaptations to better address the unique sleep challenges faced by individuals with depression.

### 4.1 Strengths and Limitations

We used a comprehensive battery of validated neuropsychological tests to objectively assess cognitive performance across multiple domains. This granular cognitive profiling allowed us to determine specific domains that may be preferentially associated with long sleep duration. The large, community-based sample of adults provided sufficient statistical power to detect associations between sleep exposures and various cognition outcomes. Our categorization of depression, considering both symptom severity and antidepressants usage status, allowed for a more nuanced examination of how depressive symptoms and treatment status might modify the sleep-cognition relationship, contributing valuable insights to the existing body of research. These findings highlight the need for targeted longitudinal and interventional research that incorporate objective sleep measures to further investigate sleep and depression as co-occurrent modifiable risk factors for cognitive decline.

Our study has some limitations. Its cross-sectional design prevented us from determining causality of the relationships we observed between sleep and cognition. We utilized self-reported measures to assess sleep duration, which can be subject to recall biases, including overestimation of actual time asleep versus time spent in bed. Additionally, some antidepressant medications can prolong sleep time and alter sleep patterns and architecture, potentially contributing to the poorer cognitive performance observed in individuals with long sleep duration and active depression.^31^ Finally, the sample primarily consisted of White participants, which limits the generalizability of the findings to more diverse populations. Longitudinal studies including diverse and ethnic backgrounds may enhance the applicability of findings to broader communities.

## 5 CONCLUSION

We found that long but not short sleep duration was associated with poorer global cognition and specific cognitive abilities like memory, visuospatial skills, and executive functions. These associations were notably stronger in people with depressive symptoms, regardless of antidepressants usage. While our study corroborates the view of the multifactorial and complex nature of cognitive function, future longitudinal studies including large-scale, multi-modal approaches—are needed to further elucidate the temporal relationship between sleep disturbances and cognitive changes. These research efforts can inform the adaptation of existing interventions to preserve cognitive health and independence in aging populations, particularly among those with depressive symptoms.

## Data Availability

Data are available on request for bone fide investigators from managing institution of the Framingham Heart Study. https://www.framinghamheartstudy.org

## Acknowledgments

We thank the research team at Boston University and all Framingham Heart Study participants who volunteered their time and made this scientific effort possible.

