## Supplementary table 1 for "Long Sleep Duration, Cognitive Performance, and the Moderating Role of Depression: A Cross-Sectional Analysis in the Framingham Heart Study"

**Supplementary table 1.** Sample characteristics by age group

| | Whole Sample | ≤44 years | ≥ 45 years |
|---|---|---|---|
| N, n (%) | 1853 | 524 (28.9) | 1329 (71.7) |
| Age at NP, years | 49.84 (9.2) | 38.77 (4.4) | 54.20 (6.5) |
| Male, n (%) | 791 (42.7) | 192 (36.6) | 599(45.1) |
| Ethnicity, n (%) | | | |
| White | 1760 (95.3) | 499 (95.2) | 1261(95.2) |
| Black | 19 (1.0) | 3 (0.6) | 16 (1.2) |
| Hispanic | 33 (1.8) | 13 (2.5) | 20 (1.5) |
| Other | 35 (1.9) | 8 (1.5) | 27 (2.0) |
| Education, n (%) | | | |
| Up to High School | 262 (14.1) | 51 (9.7) | 211 (15.9) |
| Some College | 532 (28.7) | 108 (20.6) | 424 (32.0) |
| College degree | 1059 (57.2) | 365 (69.7) | 694 (52.2) |
| Time interval [a], years | 1.69 (1.0) | 1.58 (0.9) | 1.74 (1.0) |
| APOE ε4 carriers, n (%) | 384 (21.8) | 118 (23.65) | 266 (21.0) |
| Total Cholesterol, mg/dL | 185.98 (35.0) | 180.4 (36.2) | 188.17 (34.3) |
| HDL, mg/dL | 60.53 (17.9) | 59.57 (16.2) | 60.9 (18.6) |
| Systolic BP, mm/Hg | 116 (14.1) | 111 (11.6) | 118 (14.5) |
| HTN treatment, n (%) | 390 (21.1) | 35 (6.7) | 355 (26.9) |
| Stage 1 Hypertension, n (%) | 473 (25.7) | 48 (9.2) | 425 (32.2) |
| Triglycerides, mg/dL | 112.8 (85.8) | 108.63 (95.3) | 114.44 (81.7) |
| CRP, mg/dL | 2.83 (4.5) | 3.10 (5.5) | 2.72 (4.1) |
| Body Mass Index | 28.19 (5.8) | 27.12 (5.4) | 28.61 (5.9) |
| Depression status, n (%) | 448 (24.2) | 125 (23.9) | 323 (24.3) |
| Antidepressants usage | 315 (17.0) | 79 (15.1) | 236 (17.8) |
| Depression, CES-D ≥16 | 198 (10.7) | 62 (11.8) | 136 (10.2) |
| Control [b] | 1405 (75.8) | 399 (76.2) | 1006 (75.7) |
| Antidepressants usage/CES-D <16[c] | 250 (13.5) | 63 (12.0) | 187 (14.1) |
| No Antidepressants usage/CES-D ≥ 16[d] | 133 (7.2) | 46 (8.8) | 87 (6.6) |
| Antidepressants usage/CES-D ≥16[e] | 65 (3.5) | 16 (3.1) | 49 (3.7) |

Abbreviations: NP= Neuropsychological testing, BP = Blood Pressure; HDL = high-density lipoprotein cholesterol; HTN = hypertension; CRP = C reactive protein. All values represent mean (SD) unless otherwise indicated by n (%).

NOTE. [a] time interval between self-reported sleep duration and cognitive tests. Depression groups: [b] Control = no antidepressants usage and no depressive symptoms (CES-D < 16); [c] antidepressant usage without depressive symptoms (CES-D < 16); [d] no antidepressant usage with depressive symptoms (CES-D ≥ 16); [e] antidepressant usage and with depressive symptoms (CES-D ≥ 16).
