## Supplementary table 2 for "Long Sleep Duration, Cognitive Performance, and the Moderating Role of Depression: A Cross-Sectional Analysis in the Framingham Heart Study"

**Supplementary table 2**. Cognitive characteristics by age group

| | Whole Sample | ≤44 years | ≥ 45 years |
|---|---|---|---|
| N, n (%) | 1853 | 524 (28.28) | 1329 (71.72) |
| Global Cognition [a], median [Q1, Q3] | 0.45 [-0.12,0.98] | 0.75 [0.15,1.17] | 0.34 [-0.23,0.88] |
| Trails Part A, min, median [Q1, Q3] | 0.40 [0.33,0.48] | 0.35 [0.30,0.43] | 0.42 [0.35,0.52] |
| Trails Part B, min, median [Q1, Q3] | 0.97 [0.77,1.22] | 0.85 [0.68,1.05] | 1.00 [0.80,1.28] |
| Visual Reproduction [b], n correct | 17.99 (4.89) | 19.75 (4.40) | 17.30 (4.90) |
| Logical Memories [b], n correct | 24.46 (6.74) | 25.46 (6.92) | 24.07 (6.63) |
| Similarities Test, n correct | 17.20 (3.15) | 17.28 (2.96) | 17.17 (3.22) |

Abbreviations: Min = minutes. NOTE. [a] weighted score units; [b] Sum of immediate and delayed recall scores. All values represent mean (SD) unless otherwise indicated by median [Q1, Q3].
