## Supplementary table 3 for "Long Sleep Duration, Cognitive Performance, and the Moderating Role of Depression: A Cross-Sectional Analysis in the Framingham Heart Study"

**Supplementary table 3.** Association between short and long sleep duration and cognitive scores after excluding participants < 45 years old (sensitivity analysis)

| | | Sleep Duration Categories | | | | |
|---|---|---|---|---|---|---|
| | | **Average (>6-<9 hours)** | **Short Sleep (≤6 hours)** | | **Long Sleep (≥ 9 hours)** | |
| **Cognition** | **Model** | | **β±SE** | **p** | **β±SE** | **p** |
| Global Cognition | 1 | REF | -0.03±0.05 | 0.520 | **-0.24±0.08** | **0.004** |
| | 2 | REF | -0.05±0.05 | 0.305 | **-0.27±0.09** | **0.002** |
| Trails Part A | 1 | REF | -0.009±0.02 | 0.622 | -0.02±0.03 | 0.469 |
| | 2 | REF | -0.02±0.02 | 0.273 | -0.02±0.04 | 0.566 |
| Trails Part B | 1 | REF | -0.04±0.02 | 0.099 | **-0.08±0.04** | **0.039** |
| | 2 | REF | -0.06±0.02 | 0.012 | -0.06±0.04 | 0.179 |
| Visual Reproduction | 1 | REF | -0.50±0.30 | 0.090 | **-2.05±0.52** | **<0.001** |
| | 2 | REF | -0.53±0.30 | 0.084 | **-2.14±0.56** | **<0.001** |
| Logical Memory | 1 | REF | 0.44±0.40 | 0.279 | -1.24±0.72 | 0.087 |
| | 2 | REF | 0.434±0.41 | 0.406 | **-2.20±0.76** | **0.004** |
| Similarities Test | 1 | REF | 0.09±0.19 | 0.661 | 0.03±0.35 | 0.925 |
| | 2 | REF | 0.11±0.20 | 0.594 | -0.02±0.37 | 0.949 |

NOTE. Model 1 was adjusted for age at neuropsychological testing administration, age squared at neuropsychological testing administration, education, sex, time between sleep duration questionnaires and neuropsychological testing administration and cohort. Model 2 was further adjusted for hypertension, total cholesterol level to HDL ratio, triglycerides, C reactive protein level, body max index, and APOE ε4 status. Significant findings at p <0.05 are shown in bold.
