## Supplementary table 4 for "Long Sleep Duration, Cognitive Performance, and the Moderating Role of Depression: A Cross-Sectional Analysis in the Framingham Heart Study"

**Table 4** Effect Modification by depression status on the association between self-reported sleep duration and cognitive scores after excluding participants <45 years old.

| | | Global Cognition | | Trails Part B | | Visual Reproduction | | Similarities | |
|---|---|---|---|---|---|---|---|---|---|
| | **Sleep Duration Interaction** | **p=0.009** | | **p= 0.038** | | **p=0.050** | | **p=0.006** | |
| | | **β±SE** | **p** | **β±SE** | **p** | **β±SE** | **p** | **β±SE** | **p** |
| **Control [a]** | **Average Sleep** | REF | | REF | | REF | | REF | |
| | **Short Sleep** | -0.02±0.05 | 0.714 | -0.03±0.02 | 0.203 | -0.43±0.33 | 0.195 | 0.13±0.22 | 0.543 |
| | **Long Sleep** | -0.15±0.10 | 0.157 | -0.04±0.05 | 0.481 | **-1.43±0.68** | **0.037** | 0.46±0.45 | 0.302 |
| **Antidepressants usage and CES-D <16 [b]** | Average Sleep | REF | | REF | | REF | | REF | |
| | Short Sleep | -0.04±0.14 | 0.796 | -0.09±0.07 | 0.235 | -0.56±0.86 | 0.517 | -0.12±0.56 | 0.827 |
| | Long Sleep | -0.15±0.16 | 0.369 | -0.07±0.08 | 0.391 | -1.15±0.96 | 0.236 | 0.04±0.63 | 0.950 |
| **No Antidepressants usage and CES-D ≥ 16 [c]** | Average Sleep | REF | | REF | | REF | | REF | |
| | Short Sleep | -0.22±0.17 | 0.206 | -0.12±0.09 | 0.182 | -1.62±1.20 | 0.180 | -0.18±0.68 | 0.786 |
| | Long Sleep | -0.38±0.77 | 0.623 | -0.28±0.40 | 0.484 | -5.01±5.39 | 0.356 | 0.45±3.06 | 0.884 |
| **Antidepressants usage and CES-D ≥16 [d]** | Average Sleep | REF | | REF | | REF | | REF | |
| | Short Sleep | 0.41±0.30 | 0.177 | 0.14±0.13 | 0.260 | 0.43±1.88 | 0.821 | 2.05±1.41 | 0.156 |
| | Long Sleep | **-1.10±0.35** | **0.004** | **-0.36±0.15** | **0.023** | **-6.59±2.24** | **0.005** | **-3.80±1.69** | **0.030** |

NOTE. Sleep duration categories: short ≤6h; average 7-8h; long ≥9h. Depression groups: [a] Control = no antidepressants usage and no depressive symptoms (CES-D < 16); [b] antidepressant usage without depressive symptoms (CES-D <16); [c] no antidepressant usage with depressive symptoms (CES-D ≥ 16); [d] antidepressant usage and with depressive symptoms (CES-D ≥ 16).

Model 1 was adjusted for age and age squared at neuropsychological testing, education, sex, time between sleep duration questionnaires and neuropsychological testing, and cohort. Significant findings at $p < 0.10$ for interaction and at $p < 0.05$ for the stratified model are shown in bold.

No significant interaction was found with Trails Part A (p=0.138), and Logical Memories (p=0.914).
